# Multi-Criteria Ambulance Location Problem: better to treat some fast, or more in time?

**DOI:** 10.1101/2023.01.02.23284112

**Authors:** Johanna Münch, Neele Leithäuser, Michael Moos, Jennifer Werner, Guido Scherer

## Abstract

For the location optimization of several ambulance stations, we outline a multi-criteria integer program to maximize the reached population within different time bounds. This technique is based on a realistic driving time estimation for ambulances on a detailed road network. The results show that, depending on which time bound is more valued, ambulance stations are more likely to be located in urban areas or evenly distributed across the study region. We used this model for real-world studies in Southwest Germany, where several ambulance station locations were optimized simultaneously.

## 1 Introduction

Emergency Medical Service (EMS) systems are an important part of the public health care system. In the event of an emergency, the chance of survival depends on the time that elapses before an ambulance arrives (see [10]). Therefore, the response time to emergency calls, which mainly consists of the driving time from the ambulance’s current position to the emergency scene, is an important criterion for evaluating the location structure of ambulance stations.

The Ambulance Location Problem (ALP) has been a research topic for the last decades. An extensive overview of different models can be found in [1, 13]. The ALP belongs to the class of Maximum Covering Problems (MCP) (see [13]). Most models in the literature, like ours, are extensions of the Maximal Covering Location Problem (MCLP), which was first proposed by Church and ReVelle in [3]. However, the MCLP tends to locate the facilities in densely populated areas, where the demand is higher, i.e. city centers. Therefore, rural areas are often less well covered than urban areas (see [2, 9, 10]).

Our contribution is a real-world case study of an ambulance station location problem in a rural area with some medium and large cities, that we conducted for two German city councils that needed to reopen obsolete stations (potentially at new locations). For the political decision-makers, it was very important that every inhabitant has the same chance of being supplied within the guaranteed response time, i.e. that no regions are systematically disadvantaged. We demonstrate the decision space of location alternatives that balance the goals of reaching many (urban) inhabitants in a very short response time (at the expense of (fewer) rural inhabitants that have to wait much longer) and reaching many (urban and rural) inhabitants in a longer but still good response time. In any case, the whole population of the study area must be covered within the legally allowed response time. Different response time radii were considered in [7], [4], where a double coverage within the smaller radius is maximized and the larger radius is considered for feasibility reasons. We are, however, not aware of any literature where the trade-offs between two service levels with a guaranteed maximal response time has been studied as a multi-criteria model.

Fair coverage between rural and urban areas has also been discussed in [2] and recently in [8], [9], although there is little consensus on the fairness measure. The former focuses on the ALP and also considers stochastic demand, while we assume sufficient availability of ambulances. Similarly to our approach, they use the *ε*-constraint method to compute non-dominated alternatives with respect to maximum expected coverage and one of three additional objectives to measure fairness between rural and urban regions. Therefore they penalize the longest distance or uncovered (rural) demand zones. In contrast to our case study, their demand zone resolution is much coarser. In [8], Grot et al. propose an extention to MCLP and consider fairness by maximizing the coverage of the least-covered demand area (Rawlsian criterion) and/or minimizing the differences in coverage between demand areas (Gini coefficient). Unlike us, they take site capacities into account to ensure a maximum busy fraction. We neglect this because in our (mostly rural) study area there can only be one ambulance per station in any case. While we use a detailed road network with custom speed profiles (see [11]) and inspect reachability on every road segment, the above studies use a grid based approach for their case studies with Euclidean distance, which is suitable for urban areas but not accurate enough in our study region. Luo et al. [9] present a case study also proposing a multi-objective location model. They also penalize the distance of uncovered demand points, but do not merely look at the maximum distance. Instead, they consider the weighted sum of all uncovered demand points’ distances. Additionally, they explicitly model the imbalance between urban and rural coverage as a third objective, where the minimum service level may depend on rurality. Since Luo et al. do not have emergency case data, the evaluation of their approach is yet to come.

In this paper, we outline a multi-criteria model to maximize reachability in a very short response time and a longer but still good response time simultaneously (Section 2). Our results (Section 3) clearly show, that short response times favor facilities in urban areas. In contrast to that, if the reached population is maximized within a longer response time, the stations should be evenly distributed throughout the study area and may be located in rural areas. The paper ends with an outlook and possibilities for future research.

## 2 Multi-criteria integer model

We use the following sets to formulate our model. Let *E* be the set of demand points where emergencies could occur and *S* be the set of (potential) ambulance locations. We assume that there is a subset *S*^*^ ⊂ *S* of fixed ambulance stations. Further, let *w*_*e*_ for *e* ∈ *E* denote the weight of one demand point and *d*_*s,e*_ the travel time from location *s* ∈ *S* to demand point *e* ∈ *E*. We denote with *K* ∈ ℕ the number of ambulance stations to be opened. We define three time thresholds *r*_1_ *< r*_2_ *< r*_3_ where *r*_3_ is the maximum time that a demand point *e* ∈ *E* must be reachable from an open location *s* ∈ *S*. The variables are denoted as follows:

|  |  |
| --- | --- |
| $x_s \in \{0, 1\}$ | equals 1 if station $s \in S$ is open; 0 otherwise |
| $y_{s,e}^r \in \{0, 1\}$ | equals 1 if demand point $e \in E$ is assigned to station $s \in S$ within travel time $r \in \{r_1, r_2, r_3\}$; 0 otherwise |

The model is formulated in **IP 1** (Fig. 1). Constraints (1a) ensure that every demand point *e* ∈ *E* is assigned to exactly one station *s* ∈ *S*. This assignment is only possible if station *s* is open (see constraints (1b)), while constraints (1c) ensure that a demand point *e* ∈ *E* can only be assigned to a station *s* ∈ *S* for radii *r*_1_, *r*_2_ if it is also assigned for radius *r*_3_. Constraints (1d) guarantee that a demand point *e* ∈ *E* is only assigned to a station *s* ∈ *S* within time *r* ∈ *{r*_1_, *r*_2_, *r*_3_*}* if the actual travel time *d*_*s,e*_ is at most *r*. The remaining constraints ensure that the maximal number of stations is met and that fixed stations *s S*^*^ are open. Those stations are not considered to be shifted, but they might be assigned to demand points *e ∈ E* in the considered area. All variables are binary. As objective functions, we maximize the weighted sum of assigned demand points within *r*_1_ on one hand and within *r*_2_ on the other hand.

**Fig. 1:**
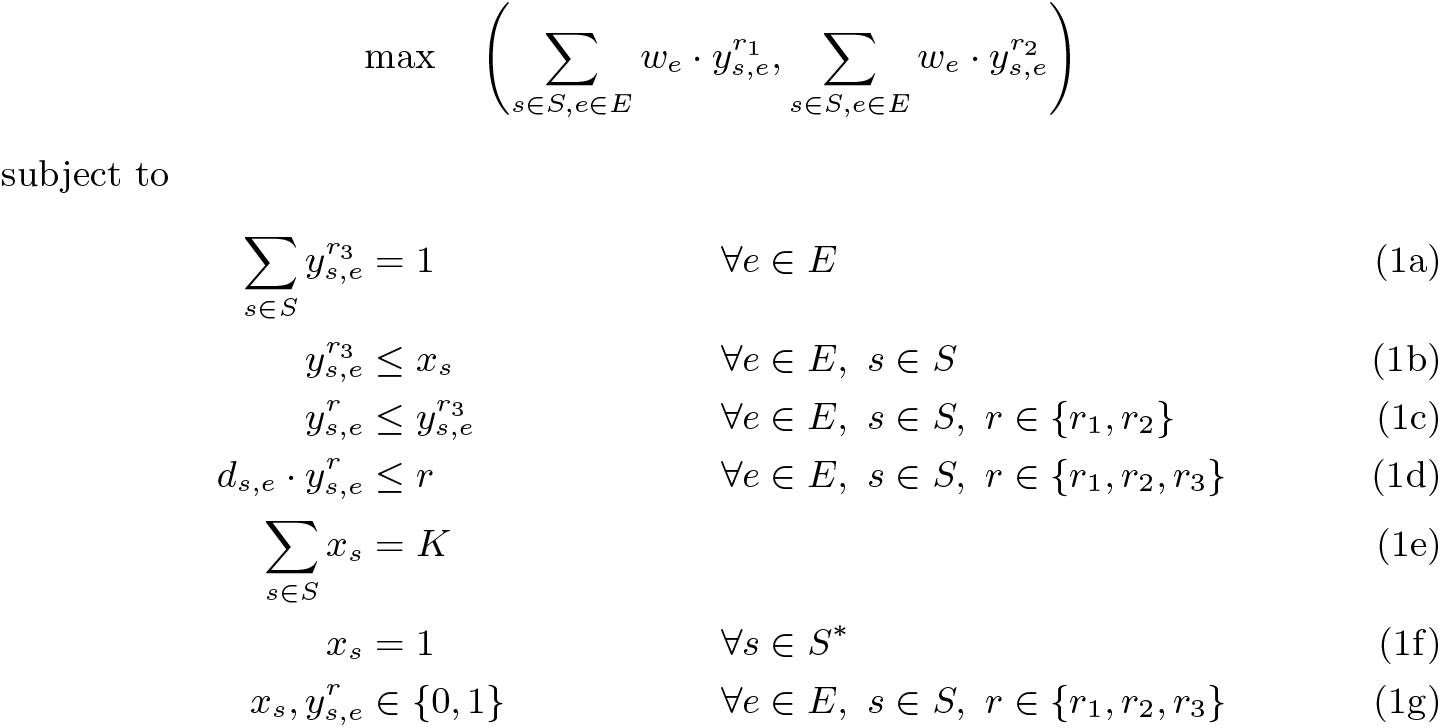
Integer program **IP 1** for the multi-criteria selection of ambulance stations.

## 3 Real-world study

We used the model **IP 1** to optimize the location of 11 outdated ambulance stations in two emergency service areas in Southwest Germany, commissioned by two district councils in Rhineland-Palatinate. Since some of these ambulance stations are directly neighboring, it was necessary to optimize the locations simultaneously. In total, we defined three study areas, which were separated from each other by fixed stations and could thus be considered individually. In each study area, three to four station locations were optimized to provide the best possible coverage with the given number of facilities. Below, we present one of the study areas exemplary.

Our client provided 17 potential locations for the four ambulance stations under consideration in this study area. Based on these 17 sites, the study area was defined to include all road sections that were fastest to reach from the potential sites. Thus, in total, the impact of the location structure on a study area with 70 municipalities, more than 764 km^2^, more than 450,000 inhabitants and more than 20,000 roads with nearly 2,300 road kilometers was studied.

Besides the 17 potential locations, from which four were to be selected, the surrounding ambulance stations in operation had to be taken into account, as there are direct interactions between the stations. Therefore, we considered in our model |*S*| = 30 ambulance stations with |*S*^*^| = 13 fixed adjacent stations. The latter stations are not yet obsolete and therefore cannot be moved. In total *K* = 13 + 4 stations must be chosen. As demand points *E*, we considered every |*E*| = 20,444 road segments and weighted them with their number of inhabitants *w*_*e*_. The travel time *d*_*s,e*_ from each station *s* ∈ *S* to each road segment *e* ∈ *E* was precalculated on a OpenStreetMap road network (graph generated with [12]). Therefore, we estimated a specific speed profile for ambulances with blue lights and siren using a large number of real emergency operations and distinguishing 29 road classes (depending on road type and maximum speed). This method was previously described in [11].

For the three different radii, we chose *r*_1_ = 5, *r*_2_ = 10 and *r*_3_ = 15. The latter one is the maximal allowed travel time in Rhineland-Palatine from an ambulance station to every public road segment. However, a response time of 7 to 8 minutes in total is medically desirable (see [10]). Since the response time also includes the call time and the turnout time, we consider a pure driving time of 5 minutes as *r*_1_. The intermediate radius *r*_2_ is chosen in between. We solved the proposed IP using Gurobi [6] within a few minutes and determine the Pareto-optimal solutions using the *ε*-constraint method described in [5, p. 98]. There are 16 Pareto-optimal solutions with respect to the population reached within 5 minutes and 10 minutes respectively. Figure 2 shows the corresponding Pareto curve.

**Fig. 2:**
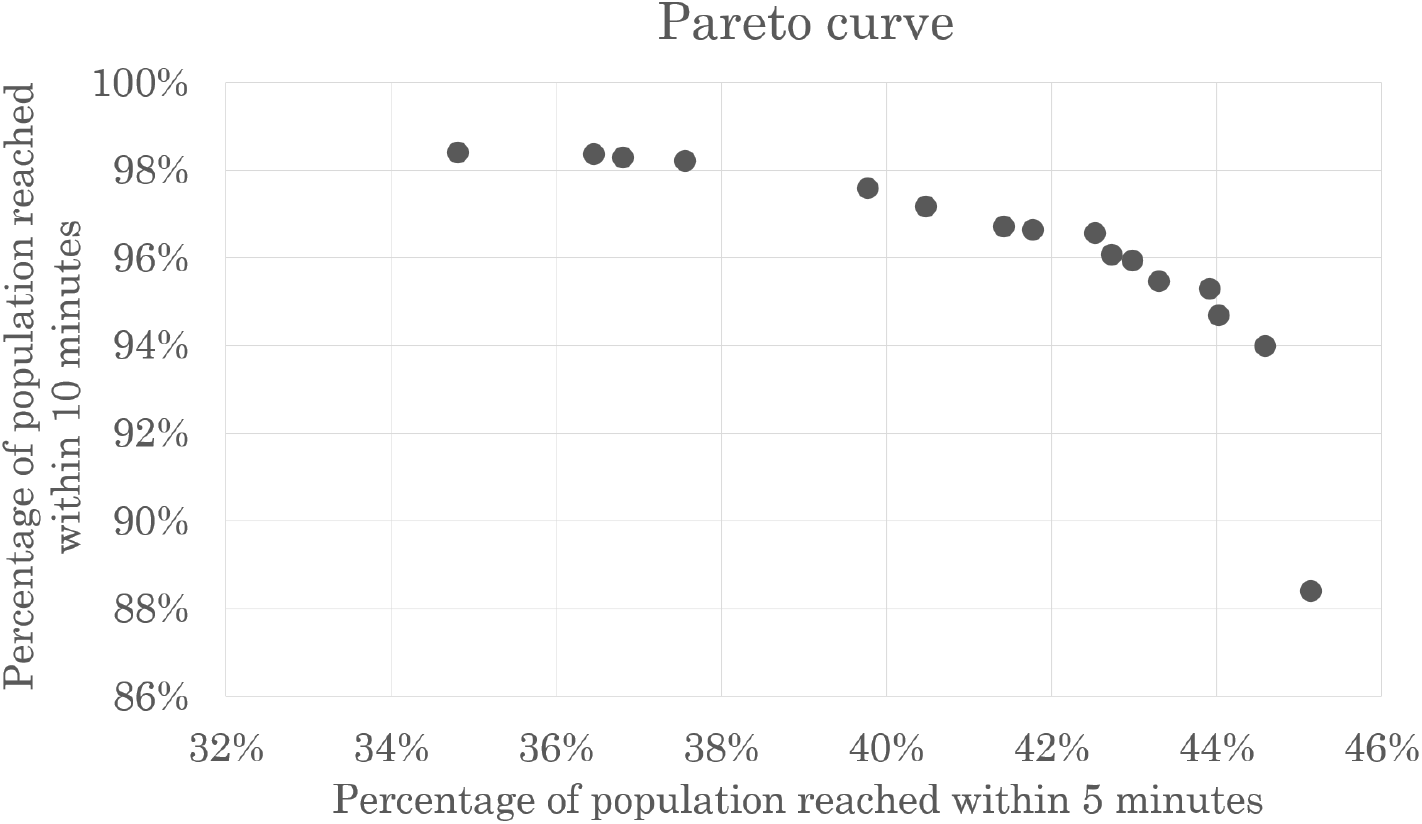
Pareto-optimal solutions with respect to the percentage of population reached within 5 and 10 minutes.

The optimization of coverage within 5 minutes clearly leads to the stations being located in urban areas where many people can be reached in a short time. If, on the other hand, the focus is exclusively on reachability in 10 minutes, the stations will be distributed much more evenly in the study area with less focus on urban populations. Depending on the geographic distribution of urban centers, these two objectives can even be completely contrary to each other.

In our case study, the contradiction becomes very clear when comparing the extreme solutions. Figure 3a shows the optimal solution maximizing the reachable population in 5 minutes. As the location combination suggests, the two largest cities are in the north and south of the study area. In opposite to that, Figure 3b shows the optimal solution maximizing the reachable population in 10 minutes. Thus, the area is covered as evenly as possible in a “zigzag” pattern and the stations are moved to mainly rural areas. However, the Pareto curve shows that starting from the extreme solutions, there is a high gain in the respective other objective possible with only a small loss in the originally single objective. Overall, in this study, the locations are pulled further and further out of the urban centers, the more value is placed on coverage within 10 minutes.

**Fig. 3:**
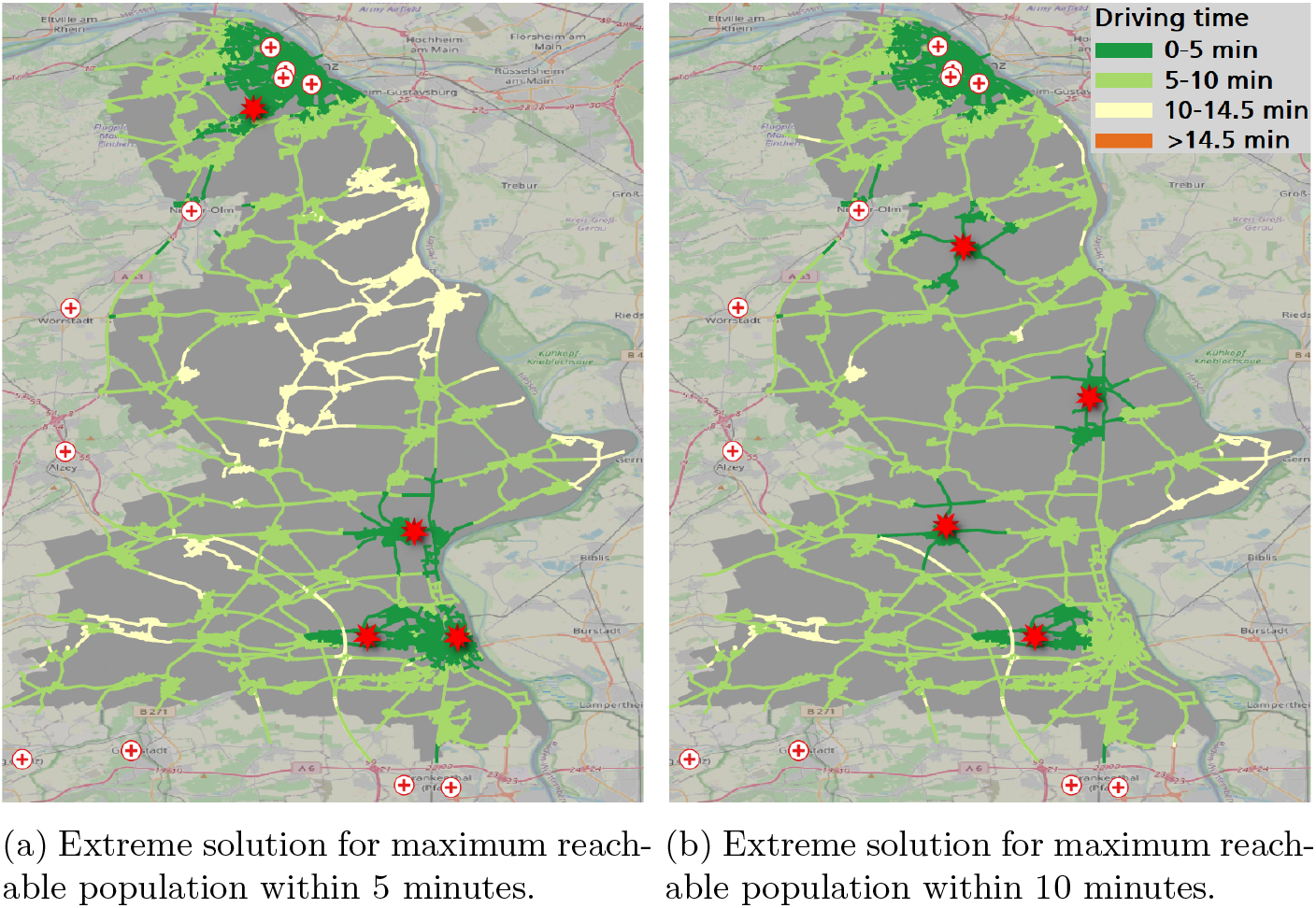
Comparison of the extreme solutions. Optimized locations are marked with a red star, fixed locations are marked with a red cross on white background. The road segments are colored according to the driving time starting from a station. The study area is colored in gray.

Using the model, we found very promising site combinations for the new stations. However, this static approach assumes that the ambulance is always present at its station when requested. Since this does not hold in reality, the identified solutions were merely used as a first stage in the decision process. They were then discussed in detail with experts from emergency medical services and the most promising solutions were further investigated using a discrete event simulation as previously described in [11]. Nevertheless, reachability in principle is a very important component for evaluating the medical care situation and legal regulations.

## 4 Conclusion and Future work

We presented a multi-criteria integer model to maximize the reachability of demand points within two radii *r*_1_ *< r*_2_, covering all demand points within *r*_3_ with *r*_3_ *> r*_2_. The model was used for a real-world study in Southwest Germany where the location of up to four ambulance stations were optimized simultaneously. As our results show, the choice of ambulance station locations depends on the weighting of the considered travel time bounds *r*_1_ and *r*_2_. If as much population as possible is to be reached in a short time for maximal survival chance in severe cases, the stations have to be located in urban areas. If, on the other hand, as much population as possible is to be reached in a longer, but legally still sufficient travel time, the stations should be distributed as evenly as possible throughout the study area and may be located in rural areas. In general the proposed model can be used for real-world studies as a decision support tool to answer the question of optimal ambulance station locations. The moral decision of placing the stations in rural or urban areas is not made by the model, but by the decision-makers.

In our study, no investment cost data were available, so the goal was to optimize the ambulance service for a given number of stations. However, our model can easily be extended to include investment costs such that the statutory minimum care can be optimized at minimal cost. At the moment, only one vehicle is planned at each of the considered stations, thus our focus was on single coverage. Nevertheless, our goal is to extend the model such that multiple coverage by different stations but also by multiple vehicles at one station is directly considered in the model. In the future, not only the location but also the provision should be optimized simultaneously depending on the population.

## Data Availability

The input data of this study are openly available.
All data produced in the present study are available upon reasonable request to the authors.

https://download.geofabrik.de/europe/germany/rheinland-pfalz.html

https://ergebnisse2011.zensus2022.de/datenbank/online/

## Notes

### Competing Interest Statement

The authors have declared no competing interest.

### Funding Statement

The results were partly obtained within the framework of a study, which was commissioned by the district administrations Bad Kreuznach and Mainz-Bingen.

